# Community-Based Culturally Tailored Education Programs for Black Adults with Cardiovascular Disease, Diabetes, Hypertension, and Stroke: A Systematic Review Protocol

**DOI:** 10.1101/2021.08.24.21262410

**Authors:** Joseph Hawkins Fulton, Hardeep Singh, Oya Pakkal, Elizabeth M Uleryk, Michelle LA Nelson

**Author notes:** **Corresponding author:** Dr. Michelle LA Nelson, 1 Bridgepoint Drive; Toronto, ON M4M 2B5.

## Abstract

**Background:** Chronic conditions and stroke disproportionately affect Black adults in communities all around the world due patterns of systemic racism, disparities in care, and lack of resources. To address unequal care received by Black communities, a shift to community-based programs that deliver culturally-tailored programs to meet the needs of the communities they serve, including Black adults who tend to have reduced access to postacute services, may give an alternative to a healthcare model which reinforces health inequities. However, community-based culturally-tailored programs (CBCT) are relatively understudied but show promise to improve the delivery of services to marginalized communities. The objectives of this review are to: (i) determine key program characteristics and outcomes of CBCT programs that are designed to improve health outcomes in Black adults with cardiovascular disease, hypertension, diabetes, or stroke and (ii) identify which of the five categories of culturally appropriate programs from Kreuter and colleagues have been used to implement CBCT programs.

**Methods:** This is a protocol for a systematic review that will search MEDLINE, EMBASE, and CINAHL databases to identify community-based culturally-tailored programs for Black adults with cardiovascular disease, hypertension, diabetes, or stroke.

**Discussion:** Health inequities have disproportionately impacted Black communities and will continue to persist if adjustments are not prioritized within healthcare to provide services, care, and programs meant to address the specific barriers to better health experienced. Many interventions meant to improve the health outcomes of marginalized groups are created with little input from target communities, leading to interventions that may not address the specific barriers contributing to poor health outcomes and are designed and implemented from an outsider’s perspective. The inclusion of community members allows for a deeper understanding of the issues facing the community and provides an opportunity to incorporate cultural values to potentially increase the efficacy, tailoring the intervention to distinct communities. An alternative to current healthcare interventions must be explored to reduce the health gap experienced by Black adults.

**Trial registration:** PROSPERO CRD42021245772

## Background

The aging population combined with medical advances have enabled people to live longer, but the proportion of people living with chronic diseases and/or with the effects of a stroke has been increasing.1 This is particularly so for Black adults (defined in this review as individuals of African descent) who experience a higher incidence and severity of stroke and their odds of cardiovascular disease, hypertension and diabetes is greater than White individuals.^2–5^ Chronic conditions such as cardiovascular disease, hypertension and diabetes not only increase the risk of additional health events, such as a stroke, but also reduce functional status, health-related quality of life, social and psychological functioning, and productivity of those living with these chronic conditions.^6^ Sharing a greater burden of these conditions has effects that extend beyond poorer physical health; these conditions can be amplified by socio-economic disparities among Black communities limiting access to the resources required for improved health.^7,8^

Numerous studies have demonstrated that Black adults receive poorer disease prevention and management care from the health system.^9–11^ For example, medical advances in identification and treatment of cardiovascular diseases have resulted in decreased incidence of and mortality from cardiovascular disease at a population-wide level. However, cardiovascular disease rates among Black adults remain high as a result of poor disease management among Black communities.^9^ Furthermore, Bonow & colleagues have found that racialized communities experience a higher frequency of undiagnosed risk factors that stem from disparities in access to care and health literacy. Black adults experience poorer health outcomes from chronic health conditions than White adults.^12,13^ Moreover, Odonkor & colleagues noted that “Black individuals were less likely to receive care that was concordant with clinical guidelines per the reported literature”.^14^

Health disparities among Black communities have been recognized for decades, but specific programs meant to address them have yet to become a priority within the medical field.^15^ Although it is not a predominant intervention strategy, a growing body of literature have called for culturally-tailored community-based (CBCT) programs to reduce health disparities and improve health outcomes for racialized communities.^15,16^ CBCT programs “provide context and meaning to the message about a given health problem or behavior” to a specific group of people based on the group’s shared “cultural values, beliefs, and behaviors”.^17^ They do so by taking into account specific factors, including social–cultural (e.g., ethnic–cultural values, racially discriminatory social policies), community (e.g., institutional racism), and familial factors (e.g., differential acculturation) that cause health disparities among specific communities.^18^ CBCT programs can be used to provide services to marginalized communities who may not receive the same levels of support from the healthcare system.^19^ In particular, they can be used to improve the health outcomes of Black Adults who have been diagnosed with hypertension and diabetes.^20–23^

Kreuter and colleagues have outlined five strategies for designing and implementing culturally tailored programs,^15^ which can serve as a valuable framework for understanding and comparing CBCT programs. The five strategies include: (1) peripheral strategies (2) evidential strategies (3) linguistic strategies (4) constituent-involving strategies and (5) sociocultural strategies.^16^ Peripheral strategies consist of creating program materials featuring images and text meant for the target cultural group. Evidential strategies employ the use of data to highlight the impacts of specific conditions on participants who are from a specific group. Linguistic strategies develop the program around the linguistic capabilities of participants using their native language or basic everyday vernacular. Constituent-involving strategies involve members of a target community to develop insights and cultural awareness in program development. Socio-cultural strategies recognize the impact of social and cultural characteristics in community health practices and leverage them to provide context into the issues facing these communities.

A common practice in CBCT programs is to involve community members. Designing CBCT programs with input and developmental insights from cultural groups that they intend to serve allows the program to target relevant barriers and challenges, allowing for resources to be applied where most needed.^15,16^ CBCT programs have shown promising impacts on chronic disease awareness, condition management, medication adherence, program satisfaction, condition specific knowledge, psychosocial factors, and health intervention strategies among Black communities.^23–28^ However, the design and structure, and operational definition of CBCT programs varies considerably from program to program, making them difficult to compare. Thus, the purpose of this systematic review is to understand the design, structure and definition of CBCT programs that have been used to improve health outcomes in Black adults with cardiovascular disease, hypertension, diabetes, or stroke using Kreuter and colleagues framework.

### Review Questions

We will address the following questions in this review:

1. What are key program characteristics and outcomes of CBCT programs that are designed to improve health outcomes in Black adults with cardiovascular disease, hypertension, diabetes, or stroke?
2. Which of the five categories of culturally appropriate programs, as identified by Kreuter and colleagues, have been used in CBCT programs for Black adults with cardiovascular disease, hypertension, diabetes, or stroke, and how have they been implemented in these programs?

## Methods

This review will follow the procedures outlined in the Preferred Reporting Items of Systematic Review of Interventions and Meta-Analyses (PRISMA) 2020 statement and has been registered with the International Prospective Register of Systematic Reviews (CRD42021245772).

### Search Strategy

To access published materials, a comprehensive search strategy will be conducted within the following electronic databases: Medline, Embase, and Cumulative Index to Nursing and Allied Health Literature (CINAHL). These particular databases were selected as they focus on health-related literature and include community-based interventions, which will allow us to locate the most relevant articles. Concepts relating to i) *community based*, ii) *culturally-tailored education*, iii) *cardiovascular disease, hypertension, diabetes* or *stroke* and iv) *Black Adults* (e.g. “African continental ancestral group”) will be searched for within the selected databases. The use of the search term “African continental ancestral group” was meant to broaden the search outside of just the American context while also recognizing Black adults as a heterogeneous population connected by a shared ancestry. A comprehensive search strategy will be created with assistance from an Information Scientist. The studies will be limited to articles published in the English-language. To enhance relevancy, we will limit to articles published on or after the year 2000. The preliminary search, which was conducted on February 24, 2021 on Medline had generated 630 articles.

### Inclusion Criteria

Detailed inclusion criteria can be found in Table 1.

**Table 1.** Inclusion Criteria.

| **Table 1. Inclusion Criteria** |  |
| --- | --- |
| **Black Adults (>18 years of age)**<br>Possible Terms:<br>-African American<br>-Black Canadian<br>-Black British<br>-Afro-Caribbean<br>-Afro-Brazilians<br>-Afro- (other countries of Central or South America) | **Other Terms for Title Abstract Screening**<br>-Minorities<br>-Visible Minority<br>-People of Color<br>-Racialized<br>-Ethnic Minority |
| **Community-Based (CB)**<br>-CB approach brings people together with the intention of sharing knowledge, experiences, and to develop a common understanding<sup>29</sup><br>-Members of the community have roles in the intervention | **Culturally-Tailored**<br>-Culturally Appropriate<br>-Culturally Competent<br>-Culturally Adapted<br>-Cultural Targeting |
| **Include at least one of the 5 Strategies of Culturally Appropriate Programs**<br>(1) peripheral strategies; (2) evidential strategies; (3) linguistic strategies; (4) constituent-involving strategies; and, (5) sociocultural strategies | **At least One Education Component**<br>Examples:<br>-Diet/ Nutrition<br>-Medication adherence<br>-Behavioral<br>-Exercise<br>-Self-Management<br>-Health Literacy<br>-Strategy (term)<br>-self-management (term) |
| **Participant has Hypertension/ Diabetes/ Stroke** | **Existing Program** |
| **Patient Focused** | **Published 2000-2021** |
| **English Studies Only** | **Study Type & Designs**<br>-Primary Source<br>-Any Empirical Study Design |

### Study design

We will include published empirical studies of any study design that gather evidence from a CBCT program for Black adults with cardiovascular disease, hypertension, diabetes, or stroke. The use of primary source studies will provide an opportunity for reviewers to extract data from the original source and ensure data specific to our topic of interest will be recorded. The inclusion of quantitative and qualitative data will allow us to generate an understanding of the effects of CBCT programs on health outcomes using quantitative data as well as patient-reported outcomes by the target community through qualitative inquiry.

### Setting

No restrictions will be placed on the country in which the program occurred. However, this review will limit to programs that take place in the community setting (i.e. outside of a hospital setting).

### Participants

The participants in this review will be Black adults (≥18 years of age) with hypertension, cardiovascular disease, diabetes, or stroke.

### Intervention

This review will include programs which deliver culturally-tailored education to the Black adults with cardiovascular disease, hypertension, diabetes, or stroke. The education should relate to the management of one of these conditions (i.e. hypertension, cardiovascular disease, diabetes, or stroke). To be considered culturally-tailored, the intervention must include at least one of the following culturally appropriate strategies outlined by Kreuter and colleagues: (1) peripheral strategies, (2) evidential strategies, (3) linguistic strategies, (4) constituent-involving strategies, and (5) sociocultural strategies.

### Type of Outcome Measure

As the intent of this review is to inform the design and structure of future programs, all outcome measurements will be reported in this systematic review, including participant-level (e.g. health-related outcomes, health literacy, medication adherence, psychosocial measurements) as well as program-level outcome measures (e.g. program adherence, satisfaction).

### Study Records

#### Search Methods

The search will be conducted by the lead author and an Information Specialist on the relevant databases. To ensure we maximize our results, we will hand search the reference lists of included studies.

#### Selection Process

All articles identified from the search will be uploaded into Covidence, a reference management software. Duplicates will be removed in Covidence. The titles and abstracts of the identified articles will be independently screened by two reviewers based on the inclusion criteria. However, prior to beginning title and abstract screening, interrater reliability with title and abstract screening among the researchers will be tested using a random sample of 50 articles. A Kappa score of ≥0.80 will be deemed acceptable as it indicates almost perfect agreement amongst the research team.^30^

Once studies are identified for the full-text screening, a similar process will be followed with two reviewers deciding if inclusion or exclusion criteria. All articles excluded at the full-text review will be categorized based on predefined terms to document the reasoning behind the exclusion. To resolve any discrepancies between reviewers during title/abstract and full-text review, meetings will be held at regular intervals to discuss disagreements and reach a team consensus.

#### Data Items and Collection Process

For each article, one researcher will complete a data extraction form to record: program names, study participants, setting, study design, duration, evaluation measure, intervention, outcome, outcome measurements and study results, to summarize the existing data.^15,31,32^ A customized data extraction form will be created based on the Joanna Briggs Institute Manual for Evidence Synthesis to identify essential program characteristics^32^ and Kreuter’s 5 strategies of culturally appropriate interventions to determine the number and type of strategies used for each program included in the review.^16^

### Quality Appraisal

Quality assessment and risk of bias will be evaluated independently by two researchers using AMSTAR: A Measurement Tool to Assess Systematic Reviews33 for all articles included in the systematic review to note the quality and detect any bias present in the study. This tool is appropriate for this review as it demonstrates substantial interrater agreement and has acceptable reliability, construct validity, and feasibility.^34^

### Data Synthesis

This systematic review of the literature will aggregate data from all studies that have used CBCT programs to improve health outcomes of Black adults with diabetes, hypertension, cardiovascular disease, or stroke using a thematic analysis.^35^ A textual description of key program characteristics (e.g. populations served, type of intervention) and outcomes of each program will be provided. Following this, an inductive thematic analysis will be used to identify similarities and differences among and within programs.^35^ A deductive thematic analysis using Kreuter and colleagues’ five categories of cultural appropriateness (i.e. peripheral, evidential, linguistic, constituent-involving, and sociocultural) will be used to determine how many, which and how these components have been used within the design and delivery of culturally-tailored education programs.^16^ These insights will inform the creation of recommendations that can guide the design and implementation of future CBCT programs for Black communities.

## Discussion

This systematic review will generate comprehensive insights into CBCT programs for Black adults with cardiovascular disease, hypertension, diabetes, or stroke. The recognition of significant health inequities within racialized communities presents a major gap in the current healthcare system where new possibilities must be explored to reduce this gap. The current healthcare system tends to work under the assumption that all ethnic/racial groups should receive the same intervention without any consideration for cultural values, beliefs, practices, customs, diets, religious views due to the programs being one-size-fits-all. CBCT programs do the opposite by acknowledging cultural differences in health experiences and are meant to address the barriers facing culturally diverse communities to improve overall health with the assistance of the community members themselves. CBCT programs are becoming more popular and more attention must be made to equity-informed intervention as valuable lessons can be learned from these programs and to apply to current and future CBCT programs.

This review will not be without limitations. First, given variability within the literature of terminology that has been used to describe CBCT programs, there is a potential we may miss relevant articles. Second, based on the heterogeneity within study designs and outcomes included in this review, we will be unable to perform a meta-analysis and therefore unable to determine the effectiveness of these programs. Third, to ensure relevancy of programs, we have restricted articles published within the last twenty years; however, we recognize that limiting the study date may exclude relevant articles published before this date. Fourth, for feasibility and resource constraints, we decided to limit inclusion of articles to those available in English; however we may miss potentially relevant literature. Lastly, to ensure we generate recommendations based on high quality evidence, we have limited this review to published articles, and as a result we will be unable to capture programs introduced within gray literature sources. Nonetheless, this review will highlight valuable insights into CBCT programs.

## Conclusion

Health inequities experienced by Black communities stem from a long history of systemic racism, differential treatment, and neglect from medical institutions. While health disparities are recognized as an issue within medicine, little has been done to shift the focus of medical research to solutions that are tailored to meet the specific needs of racialized communities. New models of care must be prioritized and researched to identify possible solutions to health inequities facing Black communities. CBCT programs do present a possible path forward to improve the health of racialized communities with messaging that is tailored to their specific cultural needs and encourages a deeper level of ownership over the type of care they receive. CBCT interventions must be assessed to determine the effectiveness of addressing health disparities in the interest.

## Data Availability

Availability of data and materials: Not Applicable

## Declarations

### Ethics approval and consent to participate

This systematic review protocol does not require ethics approval without the inclusion of human participants and will use studies who have previously obtained informed consent.

### Consent for publication

Not Applicable

### Availability of data and materials

Not Applicable

### Competing interests

The authors declare that they have no competing interests.

### Authors’ contributions

JF conceptualized the project and wrote the original protocol draft. JF, HS, and OP developed methodology, wrote, reviewed, and edited the protocol. EMU assisted in the creation of the search strategy and ran the search. MLAN contributed to the conceptualization of the project and provided supervision over the scope of the project.

## Notes

### Competing Interest Statement

The authors have declared no competing interest.

